# HIV testing by public health centers and municipalities, and new HIV cases during the COVID-19 pandemic in Japan

**DOI:** 10.1101/2020.10.16.20213959

**Authors:** Keisuke Ejima, Yoshiki Koizumi, Nao Yamamoto, Molly Rosenberg, Christina Ludema, Ana I. Bento, Daisuke Yoneoka, Seiichi Ichikawa, Daisuke Mizushima, Shingo Iwami

**Affiliations:** Department of Epidemiology and Biostatistics, Indiana University School of Public Health-Bloomington, IN, USA; Department of Global Health Policy, Graduate School of Medicine, The University of Tokyo, Tokyo, Japan; AIDS Clinical Center, National Center for Global Health and Medicine, Tokyo, Japan; School of Human Evolution and Social Change, Arizona State University, Arizona, USA; Department of Health Policy and Management, School of Medicine, Keio University, Tokyo, Japan; Graduate School of Public Health, St. Luke’s International University, Tokyo, Japan; Institute for Consumer Sciences and Human Life, Kinjo Gakuin University, Nagoya, Japan; Department of Biology, Faculty of Sciences, Kyushu University, Fukuoka, Japan; MIRAI, JST, Saitama, Japan; Institute for the Advanced Study of Human Biology (ASHBi), Kyoto University, Kyoto, Japan; NEXT-Ganken Program, Japanese Foundation for Cancer Research (JFCR), Tokyo, Japan; Science Groove Inc., Fukuoka, Japan

**Keywords:** SARS-CoV-2, COVID-19, HIV/AIDS, HIV test accessibility

## Abstract

**Background:** During the COVID-19 outbreak, medical resources were primarily allocated to COVID-19, which might have reduced facility capacity for HIV testing. Further, people may have opted against HIV testing during this period to avoid COVID-19 exposure. We investigate the influence of the COVID-19 pandemic on HIV testing and its consequences in Japan.

**Methods:** We analysed quarterly HIV/AIDS-related data from 2015 to the second quarter of 2020 using an anomaly detection approach. The data included the number of consultations that public health centers received, the number of HIV tests performed by public health centers or municipalities, and the number of newly reported HIV cases with and without AIDS diagnosis. As sensitivity analyses, we performed the same analysis for two subgroups: men who have sex with men (MSM) and non-Japanese.

**Findings:** The number of HIV tests (9,584 vs. 35,908 in the year-before period) and consultations (11,689 vs. 32,565) performed by public health centers significantly declined in the second quarter of 2020, while the proportion of HIV cases with AIDS diagnosis among all HIV cases (36·2% vs. 26·4%) significantly increased after removing the trend and seasonality effects. The number of HIV cases without AIDS diagnosis numerically decreased (166 vs. 217), although the reduction was not significant. We confirmed similar trend for the MSM and non-Japanese groups.

**Interpretation:** The current HIV testing system including public health centers misses more HIV cases at the early phase of the infection during the pandemic. Given that the clear epidemiological picture of HIV incidence during the pandemic is still uncertain, continuously monitoring the situation as well as securing sufficient test resources using self-test is essential.

**Funding:** Japan Society for the Promotion of Science, Japan Science and Technology Agency, Japan Agency for Medical Research and Development.

**Research in context:** *Evidence before this study:* Before this study, we searched PubMed, Medline, and Google Scholar on Oct 12, 2020, for articles investigated the number of HIV test and HIV cases during the COVID-19 pandemic in Japan, using the search terms “novel coronavirus” or “SARS-CoV-2”, and “HIV” or “AIDS”, and “Japan”, with no time restrictions. We found no published work relevant to our study.

*Added value of this study:* During the COVID-19 pandemic in Japan, the public health centers and municipalities temporarily suspended facility-based HIV testing to concentrate their limited resources to COVID-19 testing. We investigated the impact of the COVID-19 pandemic on the number of HIV tests in public health centers and municipalities, and on the number of HIV cases with and without AIDS diagnosis. We confirmed that the number of the test declined in the second quarter (April to June) of 2020, and the proportion of HIV with AIDS diagnosis among all HIV cases increased during the same period.

*Implications of all the available evidence:* Providing sufficient HIV testing opportunities even during the pandemic, when facility-based testing is challenging, is necessary for better clinical and public health outcomes. Self-testing and home specimen collection (e.g. dried blood spot or oral fluid test) could be a key to fill the gap between the need for HIV testing and the constraints related to the COVID-19 outbreak.

## Introduction

The ongoing COVID-19 pandemic has had broad influences on lifestyle and health issues beyond COVID-19. There have been positive impacts, such as a curtailed 2019/2020 influenza season in Japan (1), which could be partially explained by measures taken to constrain the COVID-19 outbreak. Due to reduced economic activity and human mobility during lockdown, air pollution, which is a risk factor of pulmonary diseases, was temporally improved in China (2). However, there are many problems caused by risk mitigating interventions for COVID-19, many of which are under investigation. Fear and anxiety about this new disease and changed lifestyle have been stressful for many people, which has worsened mental health (3-5).

Further, during this pandemic period, non-COVID-19 diseases have been at a lower priority for treatment because medical resources have been disproportionately allocated to treat COVID-19 patients in many health care facilities and the capacity to care for non-COVID-19 disease has been limited. Moreover, health care facilities need to balance the benefits of providing care for non-critical conditions with minimizing the risk of COVID-19 infections for both health care professionals and patients (6).

People living with HIV infections have better outcomes with early diagnosis and connection to antiretroviral treatment. Early diagnosis is also key to preventing onward transmission because early infection is characterized by high viral load and receiving an HIV diagnosis facilitates behavioural change. However, Lagat et al. reported a decline in HIV test volume in Kenya due to many barriers related to COVID-19, such as lack of funds to visit clinics and fear of COVID-19 infection at clinics (7).

To increase HIV testing opportunities, in the United States the CDC is recommending self-testing of HIV or home specimen collection (e.g. dried blood spot or oral fluid test) to avoid COVID-19 infection risk associated with face-to-face testing service (8). In Japan, the number of self-collection-based HIV tests has increased, though the majority of tests are still conducted at public health centers and clinics (i.e., facility-based testing). However, during the pandemic, public health centers and clinics have been overwhelmed by COVID-19 PCR testing and administrative work and many have temporally suspended or limited HIV testing.

AIDS is usually diagnosed by the specific symptoms and the average incubation period is 10 years (9, 10). Therefore, the impact of the pandemic on the number of reported HIV cases with AIDS diagnosis will not be substantial in a short-term period (< 1 year). In contrast, the median time interval between HIV infection to the diagnosis without AIDS symptoms is 1 ·0 years in Tokyo, Japan (11), thus the impact of shortage of the HIV test on the number of HIV cases without AIDS diagnosis will be observed even if the pandemic does not continue over a year.

We assessed temporal patterns in HIV testing during the first two quarters of 2020: 1) number of HIV consultations received by public health centers, 2) number of tests performed at public health centers and municipalities, 3) number of newly reported HIV cases without AIDS diagnosis, and 4) number of newly reported HIV cases with AIDS diagnosis. Further, we assessed whether the proportion of HIV cases with AIDS diagnosis among new HIV cases increased.

## Methods

### Data

HIV/AIDS-related data were extracted from the quarterly report from the National AIDS Surveillance Committee from the first quarter of 2015 to the second quarter of 2020 (12). The data included: (1) the number of HIV/AIDS-related consultations at public health centers, (2) the number of HIV antibody tests performed at public health centers or by municipalities, (3) the number of reported HIV cases with and without AIDS diagnosis in Japan. We further extracted the data of HIV cases with and without AIDS diagnosis for the following two subgroups: men who have sex with men (MSM) and non-Japanese, because MSM accounts for about 70% of the newly reported HIV cases (12), and foreigners are considered vulnerable during disasters due to language barriers (13). The data on the number of tests and consultations for these subgroups were not available.

Note that HIV cases with AIDS diagnosis here represent new cases of HIV identified at late-stages, compatible with an AIDS diagnosis. Thus, this number does not include HIV cases with AIDS diagnosis that were previously diagnosed and progressed to AIDS. To rephrase, the new HIV cases here are divided into two categories: early diagnosis (HIV cases without AIDS diagnoses) and late diagnosis (HIV cases with AIDS diagnoses). HIV tests at clinics/hospitals and self-testing are also available in Japan. Thus, the number of HIV testing performed by public health centers or municipalities was used to examine the trend of facility-based tests, rather than capturing the total number of HIV tests or examining the proportion of HIV positive cases among all the HIV tests.

### Anomaly detection

We applied an anomaly detection approach to those longitudinal data to identify the period when the number of HIV related consultations, HIV antibody tests, and new HIV cases with and without AIDS diagnosis were disrupted. Further, we tested when the proportion of HIV cases with AIDS diagnosis among all new HIV cases were disrupted. First, each longitudinal data was decomposed into seasonal, trend, and remainder components by seasonal-trend decomposition using LOESS (STL)(14). In STL, LOESS smoothing was iteratively used to determine the trend component. After removing the trend component, LOESS smoothing was again used to extract the seasonal component (one-year cycle). The trend component was tested whether it had an increasing or decreasing trend using a non-parametric Spearman test (15). The remainder was analyzed with an anomaly detection method, the Generalized Extreme Studentized Deviate Test (GESD), which identified outliers progressively using the deviation from mean as a test statistic (16). A statistical significance level was set at 0 ·05. All analyses were performed on the statistical computing software R 4.0.1 (R Development Core Team) with the library ‘anomalize’.

## Results

**Figure 1** shows the observed data points and the computed normal range (the shaded area). We observed a significant downward trend in HIV cases with or without AIDS diagnosis (**Figure 1AB**), all the HIV cases (**Figure 1C**), and the number of consultations (**Figure 1E**) over the period examined (from the first quarter of 2015 to the second quarter of 2020). The trend in number of tests was not significant during the period (**Figure 1D**). The proportion of HIV cases with AIDS diagnosis among all the HIV cases showed a significant downward trend during the period (**Figure 1F**), suggesting that more HIV cases have been diagnosed in the early phase of infection recently. Regarding anomaly by the last quarter of 2019 (i.e., before the COVID-19 pandemic), there are a couple of data points that significantly deviated from the normal range. However, the magnitude of the deviation of those numbers was modest.

**Figure 1.**
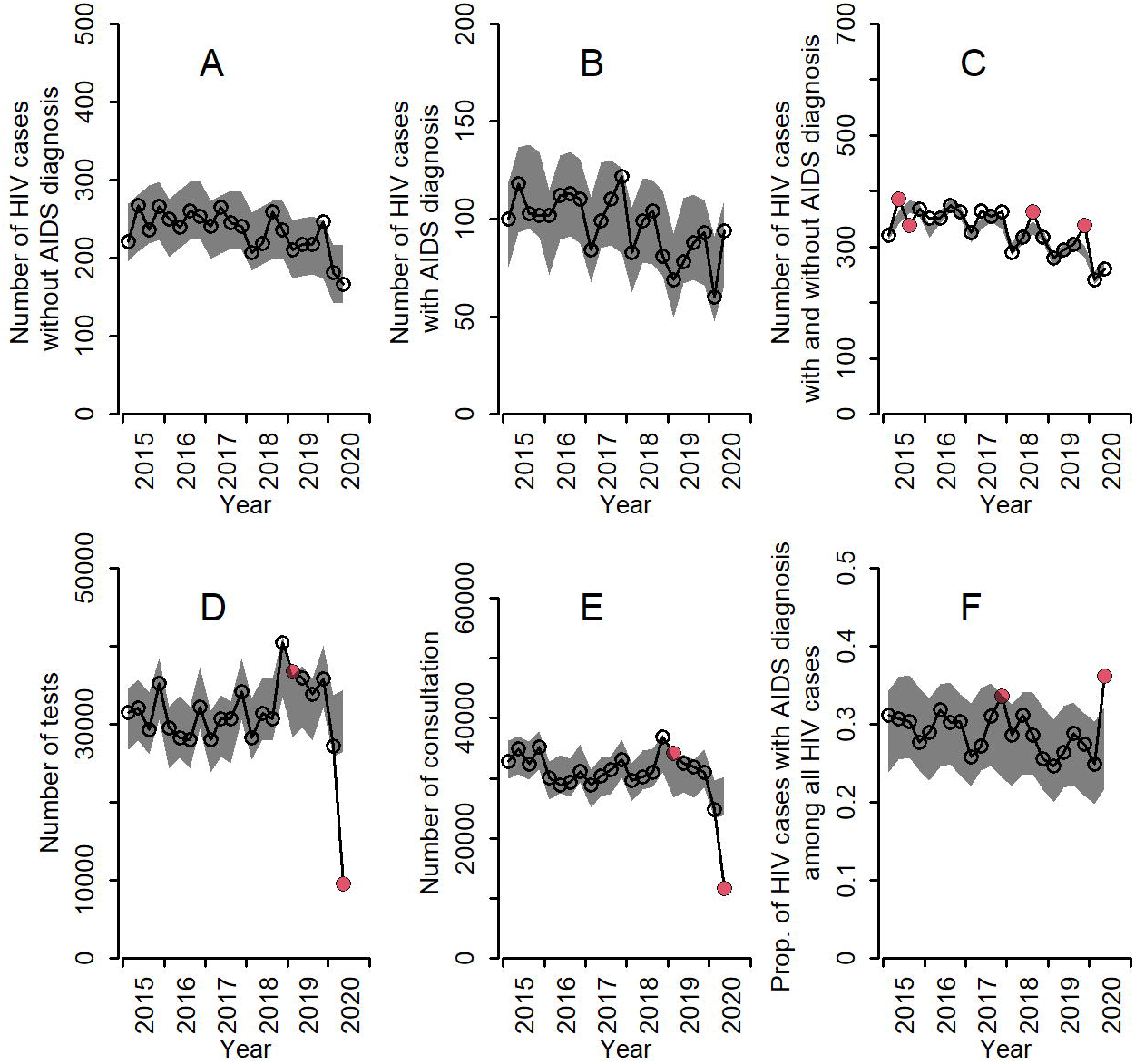
Number of HIV tests and consultations performed by public health center and municipalities, number of newly diagnosed HIV cases with and without AIDS diagnosis from 2015 to the first quarter of 2020: (A) number of newly reported HIV cases without AIDS diagnosis, (B) number of newly reported HIV cases with AIDS diagnosis, (C) number of newly reported HIV cases with and without AIDS diagnosis, (D) number of HIV test performed by public health centers or municipals, (E) number of consultation received by public health centers, (F) proportion of HIV cases with AIDS diagnosis among all positive cases. The black and red circles are observed data, and the grey shaded areas corresponds to normal range. The observed data outside of the normal rage is considered abnormal and red colored.

We did not observe deviation in any numbers in the first quarter of 2020. However, in the second quarter, the numbers of HIV tests and HIV consultations significantly declined (**Figure 1DE**), which is compatible with a public health infrastructure overwhelmed by COVID-19 related work. In contrast, the proportion of HIV cases with AIDS diagnosis among all HIV cases significantly increased (**Figure 1F**). Intriguingly, the number of HIV cases with AIDS diagnosis in the second quarter of 2020 showed the same trend as before (**Figure 1B**). The number of HIV cases without AIDS diagnosis numerically decreased in the second quarter of 2020 (**Figure 1A**), however it was not significant. It is worth noting that the number of HIV cases without AIDS diagnosis did not decline as much as the number of tests (the former dropped 23.5%, whereas the latter dropped 73.3% compared with the year-before period). This might be because more HIV cases with acute symptoms (i.e., fever) visited clinics for viral test for SARS-CoV-2 and HIV infection was subsequently diagnosed.

As a sensitivity analysis, we examined the longitudinal data of HIV cases with and without AIDS diagnosis of two subgroups: men who have sex with men (MSM) and non-Japanese. We confirmed the same downward trend for the MSM population as for the total population (**Figure 2A-C**) in terms of newly reported HIV cases with and without AIDS diagnosis. On contrary, the HIV cases without AIDS diagnosis and all HIV cases among non-Japanese were in upward trend (**Figure 3AC**). The number of HIV cases without AIDS diagnosis numerically decreased, whereas HIV cases with AIDS diagnosis numerically increased or were in the same trend as before in these two populations in the second quarter of 2020 (**Figure 2AB and 3AB**). As a result, the proportion of HIV cases with AIDS diagnosis among all positive cases also numerically increased in the second quarter of 2020 in these two subgroups (**Figure 2D and 3D**); however, they were not significant, which might be due to the small sample size. The proportion of HIV cases among MSM and non-Japanese populations relative to total HIV cases was consistent over time including the second quarter of 2020 (**Figure 2E and 3E**).

**Figure 2.**
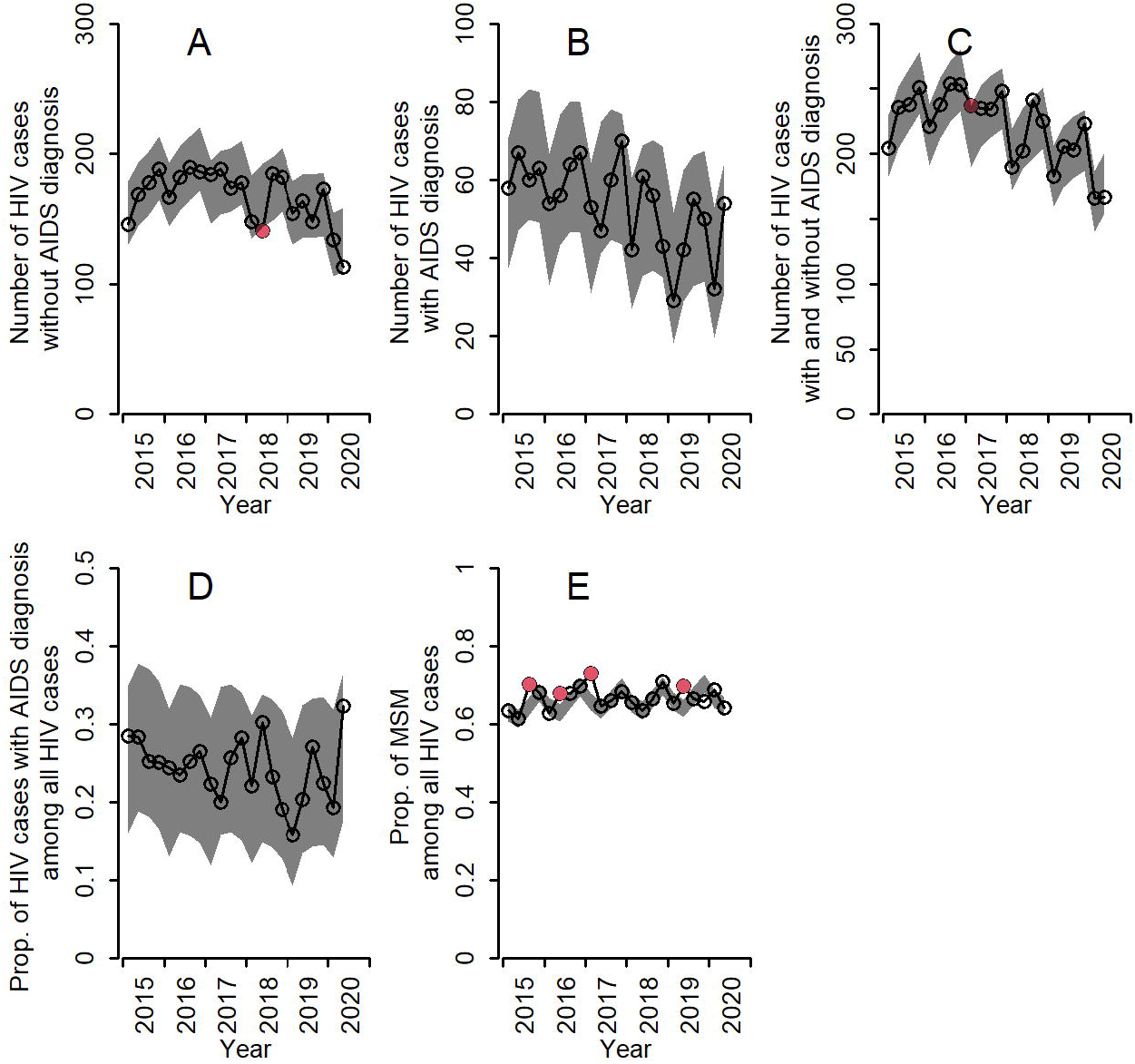
Number of newly diagnosed HIV cases with and without AIDS diagnosis for MSM from 2015 to the second quarter of 2020: (A) number of newly reported HIV cases without AIDS diagnosis, (B) number of newly reported HIV cases with AIDS diagnosis, (C) number of newly reported HIV cases with and without AIDS diagnosis, (D) proportion of HIV cases with AIDS diagnosis among all HIV cases, (E) proportion of subpopulation among all HIV cases who are MSM. The black and red circles are observed data, and the grey shaded areas corresponds to normal range. The observed data outside of the normal rage is considered abnormal and red colored.

**Figure 3.**
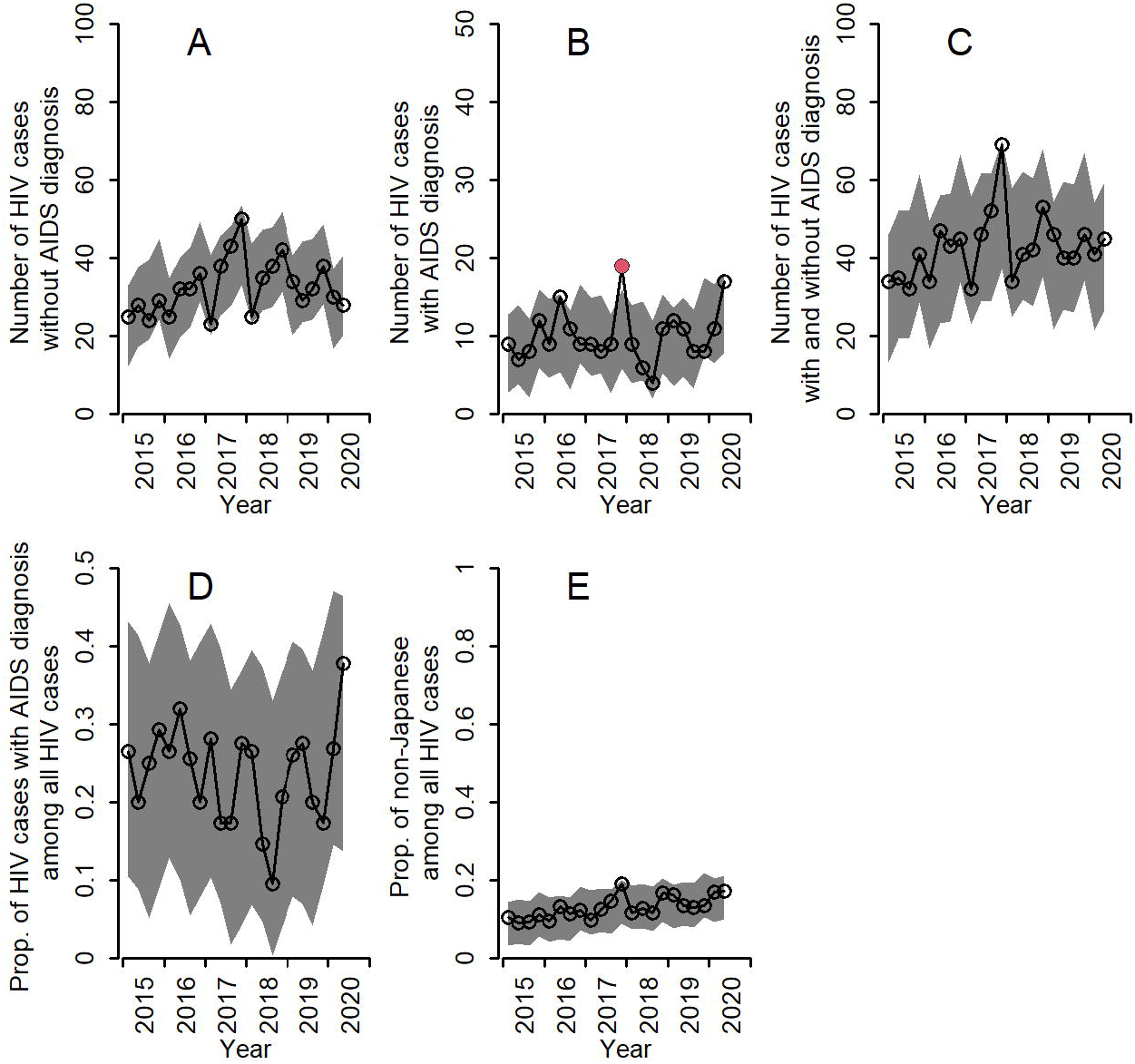
Number of newly diagnosed HIV cases with and without AIDS diagnosis for foreigners from 2015 to the second quarter of 2020: (A) number of newly reported HIV cases without AIDS diagnosis, (B) number of newly reported HIV cases with AIDS diagnosis, (C) number of newly reported HIV cases with and without AIDS diagnosis, (D) proportion of HIV cases with AIDS diagnosis among all HIV cases, (E) proportion of subpopulation among all HIV cases who are non-Japanese. The black and red circles are observed data, and the grey shaded areas corresponds to normal range. The observed data outside of the normal rage is considered abnormal and red colored.

## Discussion

A significant anomaly was identified in the number of tests and consultations performed by public health centers and municipalities, the proportion of HIV cases with AIDS diagnosis among new HIV cases in the second quarter of 2020 after removing seasonality and trend, which corresponds to the period of COVID-19 pandemic in Japan. Further, the number of HIV cases without AIDS diagnosis numerically decreased in the second quarter of 2020, although it was not significant. Under the COVID-19 pandemic situation, public health centers temporally suspended HIV tests. Further, people might have skipped the testing at public health centers because of the fear of COVID-19 infection or the state of emergency. The temporal change in the number of HIV tests, the number of HIV cases without symptoms, and the proportion of HIV cases with AIDS diagnosis may reflect such temporal change in HIV testing system in public health centers and municipalities.

We need to interpret the results carefully. First, we have analyzed the number of tests performed by public health centers and municipalities. However, this type of test accounts for less than half of the tests performed in Japan. For example, self-collection-based tests (mostly using dried blood spot) are increasing, and tests are available in clinics and hospitals; however, the count of those tests was not available. It is probable that the number of different types of HIV test has increased during the same period as compensation for the reduction in the number of tests performed by public health centers and municipalities. Second, we do not know the full picture of the impact of the pandemic on HIV epidemiology. We have used the number of reported HIV cases; however, there is likely a substantial proportion of undiagnosed cases, and it is uncertain whether the number and the proportion of undiagnosed cases changed during the pandemic. It is possible that the incidence increased (or decreased) during the pandemic. Therefore, even though the number of new HIV cases did not change dramatically, it does not indicate that the number of tests is sufficient to identify all HIV patients. Indeed, the number of reported HIV positive cases without progressed to AIDS in the second quarter of 2020 was the lowest in the last five years and the proportion of HIV cases with AIDS diagnosis significantly increased. It raises a concern that the testing opportunities might not be sufficient to fully capture the epidemiological situation of HIV/AIDS during the pandemic in Japan. Because most of the HIV cases with AIDS diagnosis are identified in clinics or hospitals due to specific symptoms, HIV cases with AIDS diagnosis are less prone to being affected by healthcare avoidance even under the pandemic. Indeed, the number of HIV cases with AIDS diagnosis was in the same trend as before the pandemic. We need to keep monitoring the situation as well as adapting testing strategies to work in these unusual circumstances.

As the pandemic continues, we do not know how long the HIV testing opportunities provided by public health centers could be disrupted and how it could affect HIV spread. Given that early detection and treatment initiation for HIV is lifesaving, providing sufficient HIV testing opportunities is important. UNAIDS has a proposed treatment target, “90-90-90” to control the HIV epidemic (17). A part of the target is that “By 2020, 90% of all people living with HIV will know their HIV status.” If the COVID-19 pandemic continuously disrupts testing opportunities, diagnosing 90% of people living with HIV might be difficult to achieve. Self-testing and home specimen collection could be a key to fill the gap between the need for HIV testing and the constraints related to the COVID-19 outbreak.

The strength of this study is that we used the quarterly HIV/AIDS reports in Japan, which include all the diagnosed cases as physicians are required to report all diagnosed cases under the Infectious Diseases Control Law(18). Further, the cases reported as HIV cases with AIDS diagnosis include only the cases of newly diagnosed HIV infection co-occurring with AIDS. Therefore, we could quantify the proportion of patients presenting with AIDS at the time of diagnosis, which captures late-presentation for diagnosis and is more likely if testing resources are not accessible.

A limitation of this study is that we were not able to account for the heterogeneous epidemiology of HIV between different regions. Higher HIV incidence has been observed in urban areas such as Tokyo and Osaka compared with rural areas. Thus, public health centers in urban areas have allocated budgetary resources towards HIV testing, whereas this has not been prioritized in rural areas. During the pandemic, the testing opportunities in rural areas may have declined even further than urban areas. Our study mainly focused on HIV cases with and without AIDS diagnosis and testing regardless of the patients’ characteristics. One of the concerns in this pandemic is that more vulnerable populations, such as sex workers, low-income populations, younger population, in addition to foreigners, may have experienced disproportionate harms. Further studies should focus on these populations and identify barriers to testing. We used only the number of tests performed by public health centers and municipalities, thus self-tests and home specimen collection, which are increasingly popular, and tests performed in clinics and hospitals were not counted. Further investigation is necessary to focus on these types of tests to fully understand the response of the HIV testing system to COVID-19 in Japan. We demonstrated that the uptake of HIV testing provided by public health centers and municipalities declined; further, HIV cases without AIDS diagnosis during the COVID-19 pandemic numerically decreased and the proportion of HIV cases with AIDS diagnosis increased. Thus, it will be critical to provide sufficient HIV testing opportunities and keep sustained attention on the unpredictable and previously unobserved public health consequences of the interactions between the dual COVID-19 and HIV epidemics.

## Data Availability

All data are publicly available from the website of National AIDS Surveillance Committee: https://api-net.jfap.or.jp/status/japan/index.html

## Author contributions

Conceived and designed the study: KE SIwami. Analysed the data: KE initially analysed the data, and NY and DY verified the analyses. Wrote the paper: KE YK NY MR CL AIB DY SIchikawa DM SIwami. All authors read and approved the final manuscript.

## Declaration of interests

The authors declare that they have no competing interests.

## Acknowledgments

This study was supported in part by Grants-in-Aid for JSPS Scientific Research (KAKENHI) Scientific Research B 18KT0018 (to S.Iwami), 18H01139 (to S.Iwami), 16H04845 (to S.Iwami), Scientific Research in Innovative Areas 20H05042 (to S.Iwami), 19H04839 (to S.Iwami), 18H05103 (to S.Iwami); AMED J-PRIDE 19fm0208006s0103 (to S.Iwami), 19fm0208014h0003 (to S.Iwami), 19fm0208019h0103 (to S.Iwami); AMED Research Program on HIV/AIDS 19fk0410023s0101 (to S.Iwami); Research Program on Emerging and Re-emerging Infectious Diseases 19fk0108050h0003 (to S.Iwami); 19fk0210036h0502 (to S.Iwami); JST MIRAI (to S.Iwami).

## Competing Interest Statement

The authors declare that they have no competing interests.

## Notes

### Competing Interest Statement

The authors have declared no competing interest.

### Author Declarations

Because we used publicly available secondary data (individuals are not identifiable), this study is not considered as a human subject study.

## References

1. Sakamoto H, Ishikane M, Ueda P. Seasonal Influenza Activity During the SARS-CoV-2 Outbreak in Japan. JAMA. 2020;323(19):1969–71.

2. He G, Pan Y, Tanaka T. The short-term impacts of COVID-19 lockdown on urban air pollution in China. Nature Sustainability. 2020.

3. Moreno C, Wykes T, Galderisi S, Nordentoft M, Crossley N, Jones N, et al. How mental health care should change as a consequence of the COVID-19 pandemic. The Lancet Psychiatry.

4. Pfefferbaum B, North CS. Mental Health and the Covid-19 Pandemic. 2020.

5. Rosenberg M, Luetke M, Hensel D, Kianersi S, Herbenick D. Depression and loneliness during COVID-19 restrictions in the United States, and their associations with frequency of social and sexual connections. 2020:2020.05.18.20101840.

6. Centers for Disease Control and Prevention. Framework for Healthcare Systems Providing Non- COVID-19 Clinical Care During the COVID-19 Pandemic 2020 [Available from: https://www.cdc.gov/coronavirus/2019-ncov/hcp/framework-non-COVID-care.html.

7. Lagat H, Sharma M, Kariithi E, Otieno G, Katz D, Masyuko S, et al. Impact of the COVID-19 Pandemic on HIV Testing and Assisted Partner Notification Services, Western Kenya. AIDS Behav. 2020:1–4.

8. Centers for Disease Control and Prevention. HIV Self Testing Guidance 2020 [Available from: https://www.cdc.gov/nchhstp/dear_colleague/2020/dcl-042820-HIV-self-testing-guidance.html.

9. Bacchetti P, Moss AR. Incubation period of AIDS in San Francisco. Nature. 1989;338(6212):251–3.

10. Muñoz A, Sabin CA, Phillips AN. The incubation period of AIDS. AIDS (London, England). 1997;11 Suppl A:S69–76.

11. Matsuoka S, Nagashima M, Sadamasu K, Mori H, Kawahata T, Zaitsu S, et al. Estimating HIV-1 incidence in Japan from the proportion of recent infections. Preventive Medicine Reports. 2019;16:100994.

12. National AIDS Surveillance Committee. Quarterly report 2020 [Available from: https://api-net.jfap.or.jp/status/japan/index.html.

13. Clark E, Fredricks K, Woc-Colburn L, Bottazzi ME, Weatherhead J. Disproportionate impact of the COVID-19 pandemic on immigrant communities in the United States. PLOS Neglected Tropical Diseases. 2020;14(7):e0008484.

14. Cleveland RB, Cleveland WS, Terpenning I. STL: A Seasonal-Trend Decomposition Procedure Based on Loess. Journal of Official Statistics. 1990;6(1):3.

15. Siegel S, Castellan N. Non parametric statistics for the behavioral sciences, Mc Graw-Mill international edition, New-York. USA; 1988.

16. Rosner B. Percentage Points for a Generalized ESD Many-Outlier Procedure. Technometrics. 1983;25(2):165–72.

17. The Joint United Nations Programme on HIV/AIDS (UNAIDS). 90-90-90: Treatment for all 2020 [Available from: https://www.unaids.org/en/resources/909090.

18. Taniguchi K, Hashimoto S, Kawado M, Murakami Y, Izumida M, Ohta A, et al. Overview of infectious disease surveillance system in Japan, 1999-2005. Journal of epidemiology. 2007;17 Suppl(Suppl):S3–S13.

